# Building and Sustaining Scholarly Communities in Health Professions Education: Lessons from U.S. Academies

**DOI:** 10.64898/2026.07.23.26358699

**Authors:** Anthony R. Artino, Zareen Zaidi, Farida Izzi, Shazia Samanani, Navin Sampathkumar, Ahdeah Pajoohesh-Ganji

**Affiliations:** Department of Health, Human Function, and Rehabilitation Sciences, School of Medicine and Health Sciences (SMHS), George Washington University (GW), Washington, DC; Division of General Internal Medicine, SMHS, GW, Washington, DC; Division of Hospital Medicine, Department of Medicine, SMHS, GW, Washington, DC; Kasturba Medical College, Manipal, India; Department of Anatomy and Cell Biology, SMHS, GW, Washington, DC

**Keywords:** health professions education scholarship, academy of educators, scholarly community of practice, faculty development infrastructure, leadership

## Abstract

**Introduction:** Academic health centers have invested in academies and related structures to promote health professions education (HPE) scholarship, yet many struggle to sustain vibrant scholarly communities. We aimed to identify lessons for building and sustaining scholarly communities that support HPE scholarship.

**Methods:** Using a constructivist approach, we conducted a qualitative interview study in 2025 with leaders from across all four geographic regions of the U.S. Inclusion criteria required that participants direct an academy or other institutional structure intended to promote educational scholarship, and that they could speak to building and sustaining scholarly communities of practice in their local context. The Communities of Practice framework informed interview guide development and data interpretation. Interviews were audio-recorded, transcribed, de-identified, and analyzed using a six-phase thematic analysis process.

**Results:** Twenty-one academy leaders described five interrelated forms of leadership work that shaped sustainable scholarly communities: (1) *building foundations that legitimize educational scholarship* by securing institutional investment, leadership buy-in, succession planning, and guiding frameworks; (2) *cultivating scholarly sanctuary to counter isolation* through psychologically safe, inclusive communities; (3) *making scholarship feasible under constraints* by creating low-barrier entry points, translating value to leaders, and addressing time, productivity, and infrastructure barriers; (4) *tending growth through layered, incremental development*, including cohort programs, consultation, mentoring, grants, and flexible participation; and (5) *making educational scholarship visible and valued* through recognition, dissemination, and outcome tracking.

**Discussion:** Viewed through the Communities of Practice framework, the themes illuminate how leaders cultivated a shared *domain* of legitimate educational scholarship, a *community* of relational support, and a *practice* of tools, routines, and developmental infrastructure. The themes also reveal that sustainable communities of practice do not emerge from any single academy model, but from context-sensitive alignment of structure, culture, and incentives. Institutions seeking to strengthen HPE scholarship should invest in foundational resources, cultivate psychologically safe communities, adopt incremental growth strategies, and create pathways for recognition and dissemination that legitimize educational scholarship.

---

Excellence in health professions education (HPE) requires more than subject matter expertise. It demands faculty who are grounded in the educational literature and scholarship, able to design instruction, assessments, and innovations informed by research and theory rather than by tradition or faculty preferences.^1,2^ Over the past two decades, academic health centers have increasingly recognized this need, responding with the creation of academies of educators, medical education research and innovation units, teaching scholars programs, and other formal structures designed to support educational excellence, innovation, and scholarly productivity.^3–7^

In this study, HPE academies serve as a primary sampling frame rather than the analytic endpoint. Our focus is on how institutions organize, resource, and sustain infrastructures that support HPE scholarship. We use the term “academy” to refer to formal membership organizations that recognize educators and provide additional benefits (e.g., faculty development) to promote teaching and educational scholarship.^5,8^ In practice, institutions also support educational scholarship through related structures (e.g., teaching scholars programs, education research and innovation units, and departments of medical education) that may not be organized as membership academies; thus, we use academy as shorthand for this broader set of scholarship-support infrastructures unless otherwise specified.

Despite institutional investment in academies and analogous support units, sustaining vibrant communities of educational scholars remains difficult. Many programs struggle with scale, succession planning, protected time, and institutional legitimacy, reflecting the broader marginalization of educational scholarship within academic medicine.^5,6,9^ Even when formal structures exist, they may function primarily as honorific bodies rather than as active engines for faculty development, collaboration, and scholarly output.^10–12^ As a result, educational scholars frequently report isolation, unclear career pathways, and limited mentorship: conditions that threaten both individual faculty trajectories and institutional aspirations for high-quality instruction, innovation, and educational scholarship.^3,13^

A growing body of literature has examined specific components of educational scholarship support, including faculty development programs focused on scholarly writing,^3,14^ mentoring initiatives,^15,16^ regional grant programs,^17^ and medical education research and innovation units.^6^ Scoping reviews and descriptive studies have identified common features associated with successful programs, such as longitudinal engagement, peer-based learning, leadership buy-in, and resource investment.^3,4,6^ However, much of this work focuses on discrete interventions or single institutional models, often emphasizing outcomes rather than the lived practices through which scholarly communities are built, led, and sustained.^5,8^ Moreover, existing studies rarely capture how academy leaders make strategic decisions about structure, culture, and growth, or navigate tensions between institutional aspirations and resource constraints.

This leaves a critical gap in the literature, which can be productively viewed through the lens of Wenger’s Communities of Practice (CoP) framework.^18,19^ The CoP framework offers a theoretical basis for examining how educational scholarship is cultivated, sustained, and legitimized within HPE academies and related institutional structures. The framework is organized around three interdependent dimensions: *domain*, the shared area of interest and collective commitment that establishes identity and purpose; *community*, the sustained relationships of mutual engagement, trust, and learning among members; and *practice*, the shared repertoire of tools, language, frameworks, methods, and ways of working that members develop and refine over time.^18,19^ Applied to HPE academies, this framework shifts attention from program outcomes to the relational, cultural, and practical processes through which leaders cultivate educational scholarship in local contexts. Guided by this framework, we conducted a qualitative study of directors of HPE academies and related institutional structures, foregrounding leaders’ experiences and decision-making to identify lessons for supporting educational scholarship and sustaining scholarly communities of practice.

## Materials and Methods

The present study employed a constructivist approach emphasizing how participants construct meaning from their experiences.^20^ We conducted semi-structured interviews to identify lessons for supporting educational scholarship and sustaining scholarly communities of HPE faculty.

### Participants and Recruitment

We collected data from February to June 2025 using purposive sampling, informed by the author team’s knowledge of individuals directing HPE academies and related structures, and further strengthened through snowball sampling. Inclusion criteria required that participants direct an academy or analogous institutional structure and have experience building and sustaining educational scholarship in their local context. Because the project was originally intended to inform the development of a local academy within a U.S. medical school, we intentionally focused recruitment on leaders at U.S. institutions. This decision reflected the importance of contextual alignment: funding models, faculty roles, promotion expectations, and organizational structures vary across international HPE systems, and we sought to maximize the practical utility of the findings for the setting in which the initiative was being developed. Within this U.S. sampling frame, we attended to institutional characteristics, including geographic region and public or private status, to achieve a balanced representation of perspectives.

Sample size was guided by the concept of information power rather than saturation.^21,22^ In this more contemporary conceptualization, sample adequacy was assessed using several criteria, including the specificity of the sample, the focus of the study aim, the quality and depth of interviews, and the extent to which the data provided sufficient richness and variation to support a credible thematic analysis.^21,22^ Given our focused aims, we estimated that a total of 20 interviews would be needed to achieve appropriate information power. After each interview, the team evaluated the quality of the data obtained, the range of perspectives included, and whether additional data might contribute new insights to the study aims.

### Ethics Statement

Ethical approval was granted by the George Washington University Office of Human Research. Prior to conducting each interview, participants received an overview of the project, and the interviews were conducted with attention to participant confidentiality and respect.

### Data Collection

We developed a semi-structured interview guide, informed by the CoP framework,^18,19^ to explore participants’ experiences with building and sustaining scholarly communities of practice in HPE. The interview guide included open-ended questions organized around the three CoP dimensions of *domain* (shared areas of interest), *community* (relationships among members), and *practice* (shared tools and resources). Each dimension was mapped to specific questions and probes, and the interview guide was designed to elicit detailed accounts of how leaders develop, support, and sustain educational scholarship.

All interviews were conducted virtually by two members of the author team (ARA and ZZ) via Zoom and lasted approximately 45-60 minutes. Demographic information was collected prior to the interviews, which were recorded and transcribed using Otter.ai, an automated, cloud-based transcription platform that uses speech recognition technology to convert spoken audio into text. Before analysis, the research team reviewed the transcripts for accuracy and completeness, made minor clarifying edits, and removed identifying information.

### Reflexivity

The research team included clinicians and PhD-trained educators from diverse professional backgrounds (e.g., education researchers, clinician-educators, a basic scientist, and a recent medical school graduate). Team members engaged in explicit reflexive dialogue through virtual meetings and email exchanges to surface assumptions about the purpose and functions of academies, as well as their own experiences with HPE scholarship. Two authors (ARA and ZZ) co-direct the Academy of Education Scholars at the George Washington University School of Medicine and Health Sciences, roles that informed their familiarity with academy structures and practices. Throughout the analysis, we reflected on how our professional roles and positionality informed interpretation.

### Data Analysis

The de-identified transcripts were uploaded into Dedoose Version 10.0.59 (SocioCultural Research Consultants, LLC, 2023), a secure online qualitative analysis platform. For analysis, we followed Braun and Clarke’s six-phase thematic analysis process to identify patterns across the dataset.^23^ In the first phase, we familiarized ourselves with the data by repeatedly reading transcripts and noting initial ideas. The second phase involved inductive open coding of transcripts, identifying features of the data that appeared meaningful in relation to the study aims, with the theoretical framework serving as a lens. The team met six times to complete the coding process. Each transcript was independently coded by two team members, and coding discrepancies were resolved through discussion during regular team meetings.

During the third phase, codes were collated into potential themes by examining how different codes combined to form overarching patterns. The fourth phase involved reviewing themes at two levels: checking that the coded extracts formed a coherent pattern within each theme and assessing whether the themes accurately reflected the meanings evident in the dataset as a whole. In the fifth phase, themes were defined and named, with ongoing analysis to refine each theme’s specifics and generate clear definitions. The final phase involved selecting compelling examples and relating the analysis back to the study aims and extant literature.

Using iterative collaborative analysis, all members of the research team reviewed, refined, and consolidated codes into themes, ensuring a comprehensive interpretation that captured the nuanced experiences of study participants. This collaborative approach enhanced the trustworthiness of findings through ongoing reflexive dialogue about analytic decisions.

## Results

We interviewed 21 academy leaders across the four geographic regions of the U.S. (central, northeast, southern, and western), as defined by the Group on Educational Affairs, Association of American Medical Colleges (see Table 1). The sample was predominantly women (81.0%), and participants were almost evenly distributed between public (52.4%) and private (47.6%) institutions; most participants held the rank of full professor (61.9%).

**Table 1.**
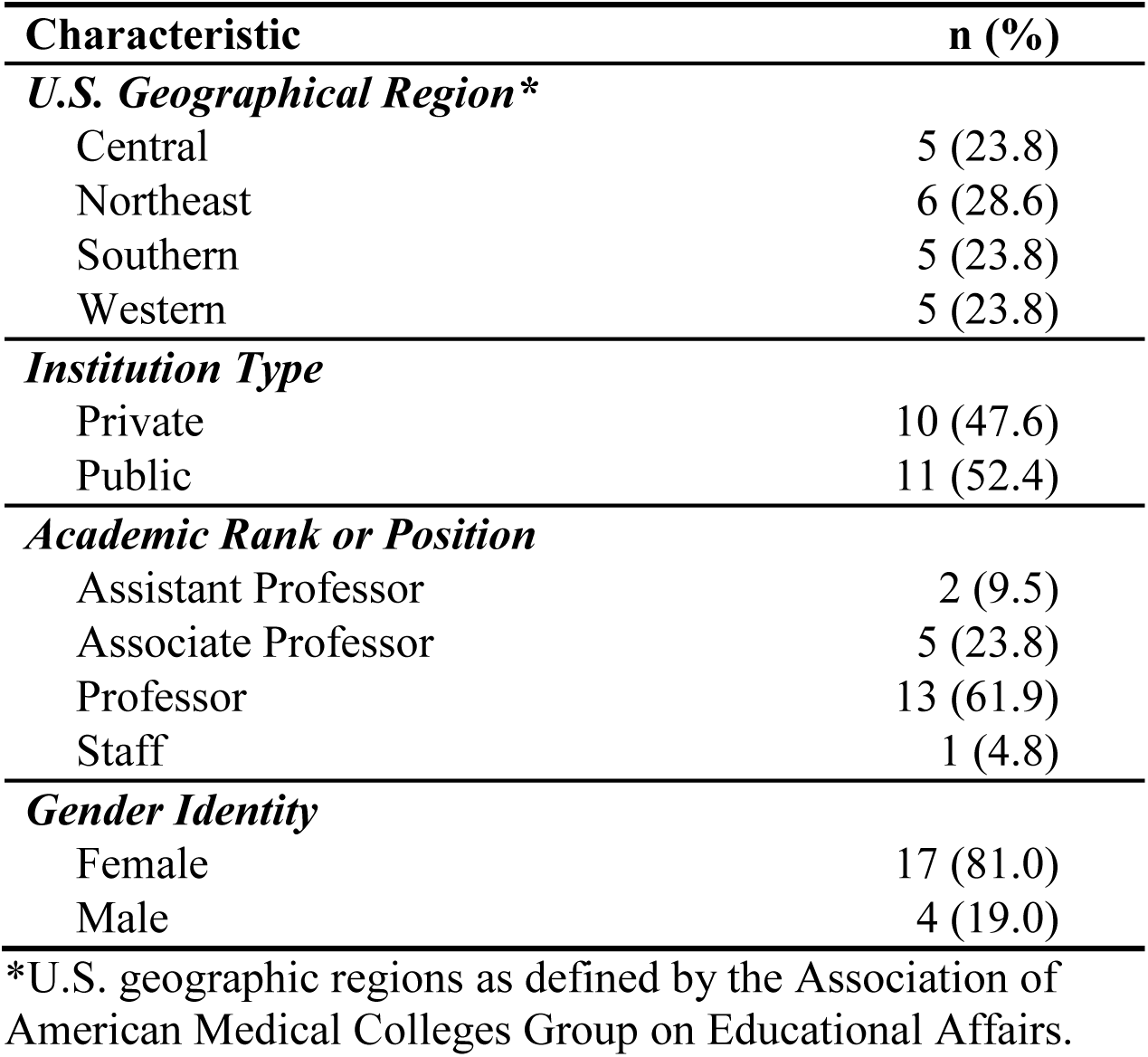
Participant Characteristics (*N* = 21)

| **Characteristic** | **n (%)** |
| --- | --- |
| ***U.S. Geographical Region\**** |  |
| Central | 5 (23.8) |
| Northeast | 6 (28.6) |
| Southern | 5 (23.8) |
| Western | 5 (23.8) |
| ***Institution Type*** |  |
| Private | 10 (47.6) |
| Public | 11 (52.4) |
| ***Academic Rank or Position*** |  |
| Assistant Professor | 2 (9.5) |
| Associate Professor | 5 (23.8) |
| Professor | 13 (61.9) |
| Staff | 1 (4.8) |
| ***Gender Identity*** |  |
| Female | 17 (81.0) |
| Male | 4 (19.0) |

\*U.S. geographic regions as defined by the Association of American Medical Colleges Group on Educational Affairs.

Across interviews, academy leaders described sustainability not as the result of a single or predominant academy model, but as the product of recurring, interrelated forms of leadership work. This work included legitimizing educational scholarship as a shared institutional domain, cultivating relational spaces where educators could engage in scholarship together, adapting to persistent structural constraints, developing scholarly capacity incrementally, and making educational scholarship visible and valued within the institution. The five themes described in detail below reflect these interrelated forms of leadership work, and the strategies leaders employed often served multiple purposes simultaneously. For example, mentoring initiatives both strengthened community and developed scholarly capacity, while dissemination activities recognized accomplishments and reinforced the institutional legitimacy of educational scholarship. Although some strategies therefore appeared across more than one theme, each theme captures the primary function those strategies served in sustaining scholarly communities of practice. Together, the themes illuminate how leaders cultivate the domain, community, and practice of HPE scholarship through overlapping but analytically distinct forms of work.

### Theme 1: Building Foundations That Legitimize Educational Scholarship

Participants described foundation-building as the work of translating educational scholarship from an individual interest or honorific identity into a legitimate institutional domain. Across sites, this required more than creating programs; it required visible leadership support, dedicated resources, succession planning, shared values, and organizing frameworks that gave educational scholarship structure and continuity. Several participants described programs that emerged from identified institutional needs or from efforts to relaunch initiatives that had become dormant or primarily symbolic. As one participant explained,

> The Academy of X existed for five or six years, but then had been dormant for four or five years… The way it worked at that point was that you sort of applied, and showed your materials you put in, you got inducted to the academy, you put it on your CV, and kind of basically you were done. (P10)

Recognizing the limitations of purely honorific structures, leaders described relaunching programs around active scholarly engagement. The need for “succession planning” (P8) was also identified as critical, with participants noting that leadership transitions without clear succession strategies led to periods of institutional dormancy and instability. Relatedly, participants emphasized the importance of shared leadership, particularly in the form of co-director dyads.

One participant reflected, “I could not have done any of this myself,” adding that the answer was not necessarily “having a committee,” but rather having “at least one other person” who either sees the world similarly or complements one’s vision in ways that allow leaders “to really build something that’s going to have broad appeal” (P2).

Institutional support emerged as another way leaders established educational scholarship as legitimate and durable, rather than peripheral or dependent on individual enthusiasm. One participant articulated the comprehensive nature of this investment,

> We’ve been very well supported… we have funding for full-time equivalents (FTE), so 50% of my job, that’s a significant investment from the school. One full-time staff member… another .6 FTE staff who supports the teaching scholars program… you’re running into very quickly, hundreds of thousands of dollars to support this program, just from a personnel perspective. (P14)

Leadership buy-in at the highest levels was also deemed essential, with participants emphasizing the need for deans and vice deans to provide not just financial resources but also administrative support, protected time, and institutional positioning. A participant described receiving a “blank sheet of paper” (P13) from institutional leaders (i.e., a blank check or blank slate), signifying both trust and a mandate to build their program from the ground up.

Participants described how guiding frameworks and core values gave programs coherence. They referenced models such as “Kirkpatrick” (P19) for evaluation, as well as institutional frameworks (P8) for organizing educational domains. The diversity of approaches was captured by one participant’s observation that “if you see one academy, you see one academy” (P19), underscoring the need for contextual adaptation while maintaining theoretical grounding.

Programs were organized around clear missions, typically encompassing “faculty development,” “recognition,” and “research” (P13), and around core values such as “excellence,” “support,” and “generosity” (P1). Tiered membership structures, ranging from “associate member” to “distinguished educator” (P9), created pathways for progressive engagement and also helped to align recognition with institutional promotion processes. Together, these structures, values, and frameworks helped define the domain of educational scholarship and signal that it counted within the institution.

### Theme 2: Cultivating Scholarly Sanctuary to Counter Isolation

Participants described sustainable scholarly communities as relational spaces that countered isolation and made educational scholarship feel safe, shared, and possible. In this sense, sanctuary was not simply collegiality; it was a deliberate leadership strategy for lowering status barriers, supporting unfinished scholarly work, and helping educators learn across disciplinary boundaries. The “community of practice” was a core organizing principle, with participants describing how these communities combat the isolation inherent in educational scholarship. One participant reflected: “I think that one should never be doing research alone (except as part of a dissertation process)… it’s so isolating doing this stuff independently, having a community of practice makes all the difference” (P19). Programs prioritized “community building” (P8) through multiple mechanisms, including regular gatherings, conference support, and informal networks of “champions and ambassadors” (P8) embedded within departments.

Psychological safety emerged as essential for meaningful participation, particularly given the interdisciplinary nature of educational scholarship. One participant explained the intentional cultivation of safe spaces,

> We really want to cultivate an environment that feels safe, because sometimes you get people in the room who have PhDs, and they think they’re, you know, God’s gift, and it can be really intimidating. And we really work to not have that happen. (P1)

The value of “authenticity” (P19) was also emphasized, with leaders modeling vulnerability to encourage participants to “drop their guard” (P19) and engage openly with educational works in progress.

Participants described building inclusive community-oriented structures. One participant described their initial resistance to the idea of an academy, explaining, “I didn’t want an academy because I felt like academies can be exclusive as opposed to inclusive” (P7). This concern shifted as the academy took shape as an intentionally inclusive structure. Programs emphasized being “built by the members, for the members” (P21), with governance structures that prioritized bottom-up rather than top-down approaches. This inclusive orientation extended to diverse audiences, with programs serving “MDs,” “PAs,” and “PhDs” (P12), and hosting events open to the broader institutional community. Some programs strategically focused on “niche” communities of committed scholars (P1), while others cast wider nets to maximize engagement, reflecting varied approaches to balancing depth and breadth of community building. The cultivation of a “culture of scholarship” (P8) where “everything we do, we do in a scholarly way” (P8) represented the aspirational endpoint for many of these community-building efforts.

### Theme 3: Making Scholarship Feasible Under Constraints

Participants emphasized that they were rarely able to completely eliminate the structural barriers facing educational scholars. Instead, they made scholarship more feasible by designing low-barrier entry points, translating educational scholarship into institutionally legible value, and creating pragmatic supports that allowed scholarly work to continue under pressure.

Unsurprisingly, lack of time and clinical pressures emerged as the most pervasive challenges, with one participant stating simply: “I think the biggest challenge is time because everybody’s competing against a very busy clinical faculty’s time” (P12). The dominance of clinical productivity metrics created structural barriers, as participants noted that “clinical RVUs” (P12) drive compensation while “education’s always the first to go” when resources become constrained. The broader healthcare environment intensified these pressures, with one participant observing that “it’s all about production. It’s all about clinical revenue” (P11), creating an institutional context where educational scholarship struggles to gain recognition and prominence.

In response to these constraints, participants described a range of pragmatic, adaptive solutions designed to work within, rather than against, existing institutional realities. Several leaders intentionally created low-barrier scholarly entry points that minimized preparation demands while fostering connection and momentum. For example, one participant described hosting recurring discussion circles built around short blog posts, emphasizing that “you don’t have to do any pre-work. You show up, I give you lunch, and then we read the blog post… we do it all in an hour” (P7). These sessions were co-facilitated by faculty and trainees from diverse roles, making educational scholarship feel “more approachable to others who might want to know about it” (P7) while respecting intense clinical schedules.

Participants also described strategies to counter fragmented scholarly support and disciplinary silos. These silos mattered because they left educators with project ideas but without stable communities, methodological support, or shared ownership needed to carry projects forward. One leader detailed the evolution of “mentorship circles” (P6) that intentionally moved faculty from isolated, project-specific interests toward shared scholarly work. After months of difficulty encouraging collaboration, this leader formalized a small, topic-focused “lab,” explaining: “I’m making you all members of the lab, and we’re going to meet on a regular basis” (P6). By structuring recurring interaction and shared ownership, participants reframed mentorship as a collective, relational process rather than an individualized resource.

Participants noted that infrastructure gaps compounded time constraints. These gaps were not limited to staffing or administrative support; they also included a lack of shared language, methodological scaffolding, and institutional understanding of why educational scholarship mattered. They described the challenge of helping colleagues understand “what is researchable” (P4) and the exhausting reality of “building a house every single time” (P4) without established research programs to scaffold new scholars’ work. One leader reflected on persistent skepticism: “People would just raise the question, well, why do we even need this? We need teachers, yes, but we don’t really need people who study teaching” (P2). This lack of understanding of the importance of educational scholarship extended to institutional leadership. In response, several leaders emphasized the importance of advocacy and translation: developing “compelling reports” (P18) and narratives that resonated with institutional leaders unfamiliar with educational scholarship.

Despite the range of pragmatic, adaptive solutions described above, protected time for scholarship remained elusive for most participants. One senior faculty member reflected,

> I’ve never had funded FTE to do scholarship… Years down the road, and it’s like, I never had funded time to do the work. It was all choices I made that were built on taking on administrative roles that would give me flexibility to do scholarship around that. (P14)

Leaders’ own experiences with unfunded scholarship shaped how they designed supports for others facing similar constraints. Rather than viewing administrative work as separate from scholarship, some participants strategically leveraged these roles to create flexibility, visibility, and advocacy capacity, positioning themselves to protect and legitimize educational work within institutional decision-making structures. In addition, because buying out faculty time was costly, especially clinician time, one participant instead used funds to “pull in extenders” (P6): individuals who assisted faculty with research tasks such as literature searching and statistical analysis.

Notwithstanding these constraints, participants demonstrated persistence, with one noting, “It takes energy to keep pushing education scholarship to the forefront” (P21), acknowledging both the weight of barriers and the resolve required to overcome them.

### Theme 4: Tending Growth through Layered, Incremental Development

Participants described growth as a gradual process of layering opportunities over time rather than scaling rapidly. This deliberate, incremental approach mattered because leaders were trying to expand scholarly capacity without overwhelming faculty, diluting expertise, or outstripping available resources. Several participants cautioned against growing academies “too big too fast” (P12) or becoming “too broad” (P13) before developing focused expertise. In this context, flexible participation was not simply a convenience but a strategy for sustaining engagement over time. One leader explained,

> A lot of our educators have other activities that they need to attend to, and if you disrespect that, that’s when you lose them. If you show that you care about them and that there’s enough variability in timing and in options and offerings that they can participate in, that will be helpful. (P13)

Many programs were developed over extended timeframes, with one participant noting a “10-year establishment” (P21) period before achieving stability. Capacity-building, rather than one-time programming, was central to this growth. Leaders noted that education and training in educational research methods and theory were essential, despite common misconceptions. As one participant noted: “It does take expertise to do education scholarship, even though there’s a fair number of people who think anyone can do it” (P8). Programs developed diverse educational offerings, including monthly “brown bag” sessions with “theory corner” discussions and “works in progress presentations” (P8), leadership development programs for “mid-career educators” (P13), and structured mentoring initiatives. Mentoring models varied, with some emphasizing departmental approaches where “senior faculty or mid-career faculty working with a junior faculty on a project together” proved most effective (P12), while others created “mentoring pods” (P19) to connect scholars with similar interests into collaborative groups. These offerings served as growth infrastructure, helping participants move from interest to shared scholarly practice.

Program leaders balanced structured curricula with responsive consultation services. One participant articulated this layered approach,

> I really think a cohort program is very valuable. I think it builds the community of practice instantaneously… But I think you need to layer it. I think you need to have the consultation option and need to have some little entry-level faculty development opportunities, and I do think having support from, you know, a grants opportunity and that kind of thing is also really crucial. (P16)

The meeting formats used by various leaders evolved to accommodate geographic dispersion, with programs using “blended virtual/in-person” approaches (P21) and responsive scheduling.

Leaders maintained visions for sustainable growth, including incorporating program “graduates as mentors for the next generation” (P16) and establishing endowed resources to “crystallize” programming ideas (P8).

### Theme 5: Making Educational Scholarship Visible and Valued

Participants described visibility as essential to sustainability. Recognition, grants, dissemination platforms, promotion signals, and outcome tracking did more than showcase accomplishments; they translated educational scholarship into forms of institutional value. Internal (or intramural) grant programs were described as crucial for amplifying educational research and innovation projects, providing necessary seed funding. Some institutions described small awards ranging from “$4,000 to $5,000 small grants” (P14) for specific administrative needs, while others discussed substantial “$100,000” annual programs (P16). One participant explained,

> The core piece of what we do is a small grants program. This is a competitive, small grant program. This year, we had 29 applicants, and we ended up funding 6 applicants… A number of folks have published their work using support from the small grants program. (P14)

Travel grants supported conference attendance for “emerging scholars” (P21), while aspire grants provided targeted support for “biostatistics, executive coaching,” and “editorial support” (P13).

Some programs made educational scholarship visible by creating platforms for dissemination at multiple levels. Institutional events like “annual teaching and learning symposiums” (P17) and “educational scholarship days” (P21) showcased faculty work, while “writing camps” produced tangible outputs, with one program yielding “five publications from that group after a course of six weeks” (P13). External opportunities expanded reach, with some programs facilitating placements in prestigious fellowships such as “Harvard Macy” (P20) through institutional vetting processes. Leaders recognized the importance of making scholarship “real” by connecting it to “quality improvement” and “patient care” (P13), demonstrating the tangible impact of educational scholarship beyond peer-reviewed publications.

Outcome tracking provided another form of visibility by making educational scholarship legible to institutional audiences who might otherwise overlook it. Tracking and reporting outcomes helped demonstrate value, guide program development, and make educational scholarship more visible. Participants described systems for monitoring “50-60% participation rates” (P21), publications, presentations, and attainment of leadership roles. The connection between academy membership and promotion emerged as particularly significant: “When the promotions committee sees that someone is a member of the teaching academy, the conversation kind of pauses like, oh, well, their education credentials have already been vetted by the academy… that means something” (P9). Programs cultivated broader visibility through institutional newsletters (P6) and social media engagement. One participant noted that an article had “20,000 views,” which counted toward promotion (P13), and another described monthly blog posts authored by “faculty, staff, and trainees” (P7). Participants described advocacy efforts on institutional committees (P13), “road show presentations” (P3) about scholarly approaches to teaching, and rehearsal sessions before conferences (P8). Through these varied mechanisms, programs worked to establish educational scholarship as visible, valued, and viable within their institutions.

## Discussion

In this qualitative study of leaders of academies and related scholarship-support structures, we identified five interrelated forms of leadership work that illuminate how institutions build and sustain scholarly communities in HPE: (1) *building foundations that legitimize educational scholarship*, (2) *cultivating scholarly sanctuary to counter isolation*, (3) *making scholarship feasible under constraints*, (4) *tending growth through layered, incremental development*, and (5) *making educational scholarship visible and valued*. When viewed through the CoP framework, these five themes describe the institutional, relational, and practical conditions that shape the three dimensions of domain, community, and practice. *Domain* (the legitimacy and shared purpose of educational scholarship) was established through foundational investments, explicit values and frameworks, and recognition systems that signaled educational scholarship as a valued area of inquiry (themes 1 and 5). *Community* (the relational fabric of mutual engagement and trust) was cultivated through psychologically safe, inclusive sanctuaries (theme 2) and sustained through incremental developmental structures that allowed educators across career stages and disciplines to engage with one another (theme 4). *Practice* (the shared repertoire of methods, language, tools, and ways of working) was developed as leaders created pragmatic supports under constraints (theme 3) and layered programs, consultation, mentoring, grants, and dissemination opportunities over time (themes 4 and 5).

Taken together, these findings reinforce a central insight captured by participants themselves: there is no single academy model that guarantees a sustainable community of practice. Rather, sustainable scholarly communities emerge from deliberate, context-sensitive alignment of structure, culture, and institutional incentives. In this study, participants described sustainability not as a fixed endpoint, but as the ongoing alignment of shared purpose, meaningful relationships, and supported scholarly activity: the core dimensions of domain, community, and practice within the CoP framework.

Our findings extend and deepen prior descriptive and scoping work on academies, medical education research and innovation units, and faculty development programs focused on educational scholarship. Previous studies have emphasized the importance of leadership buy-in and advocacy, longitudinal engagement, mentoring, and resource investment as core features of successful educational scholarship initiatives.^3–6,24^ Our study corroborates these elements and illustrates how leaders operationalize them in practice.

Consistent with prior critiques of honorific academies,^12^ participants described a deliberate shift away from static recognition models toward structures that function as active engines of faculty development and scholarly production.^4,25^ Tiered membership, explicit missions linking faculty development, research, and recognition, and intentional succession planning were described as mechanisms for preventing program stagnation and ensuring continuity over time.

The emphasis on sanctuary (i.e., psychological safety, inclusivity, and community) adds important nuance to existing accounts of educational scholarship infrastructure. While prior work has documented isolation among clinician-educators and educational scholars,^9^ our findings demonstrate that leaders view community-building not as ancillary but as foundational. These communities of practice counter isolation, legitimize educational inquiry, and illustrate how the domain, community, and practice dimensions of the CoP framework are enacted in HPE settings.^26,27^

At the same time, participants underscored the enduring structural constraints facing educational scholars. Time scarcity, clinical productivity pressures, and limited protected time remain pervasive challenges, echoing longstanding concerns about the marginalization of education within academic medicine.^28–30^ Notably, even leaders of well-resourced programs described sustaining momentum as an ongoing act of advocacy rather than a solved problem. In response, participants emphasized the importance of pragmatic, context-sensitive strategies, such as creating low-burden scholarly entry points (e.g., “no-prep journal clubs”^31^), cultivating relational mentoring structures, and leveraging leadership roles to increase flexibility and visibility for educational work. These findings emphasize that establishing and sustaining an academy is not an endpoint but a process that requires ongoing effort to sustain visibility and viability.^4^

Across interviews, participants consistently pointed to a small set of high-leverage institutional supports that helped make scholarly participation more feasible and sustainable: dedicated and sustained funding for faculty and staff effort, including even modest FTE support; formalized mentorship structures with protected time for mentors; longitudinal, cohort-based faculty development programs that integrate project-based learning; and institutional recognition systems, such as promotion criteria and awards, that legitimize educational scholarship.

Altogether, these findings suggest that individual-level interventions, such as workshops or one-time faculty development sessions, are insufficient on their own. Meaningful progress requires alignment between faculty development efforts and broader institutional structures that provide time, mentorship, resources, and recognition for educational scholarship.

Finally, our findings about making educational scholarship visible and valued resonate with the literature emphasizing the importance of recognition, dissemination, and promotion alignment for legitimizing educational scholarship.^3,4,6^ Participants described a wide array of strategies, including internal grants, dissemination initiatives, and outcome tracking that collectively translated educational scholarship into institutional value. Importantly, these mechanisms served not only to reward individual faculty but also to communicate, internally and externally, that educational scholarship “counts.”

As a whole, these findings suggest that institutions seeking to build or revitalize academies of education scholars, or create other mechanisms for supporting educational scholarship, should resist the temptation to replicate external models wholesale. Instead, leaders might focus on articulating clear purposes, investing in people and relationships, and aligning recognition structures with institutional priorities. Sustainable scholarly communities of practice appear less dependent on any particular organizational form than on their leaders’ ability to cultivate shared purpose (*domain*), meaningful relationships (*community*), and supported scholarly activity (*practice*), and to balance inclusivity with rigor, growth with feasibility, and aspiration with available resources.^18,19^ In this sense, the observation that “if you’ve seen one academy, you’ve seen one academy” is not a weakness of the movement, but a reflection of its contextual responsiveness.

This study has several limitations. Participants were purposefully recruited through professional networks and snowball sampling, which may limit transferability, particularly to institutions with less-developed educational scholarship ecosystems. Furthermore, although our sample was limited to U.S. institutions, many of the challenges identified, such as limited protected time, the need for mentorship, and tensions in defining the role of academies, likely apply internationally. At the same time, structural differences across countries may shape how these challenges are experienced and addressed. Future work should extend this line of inquiry to non-U.S. contexts to better understand how local conditions influence the development of scholarly communities of practice in HPE. In addition, our focus on the perspectives of academy directors and related scholarship-support structures also means that participants’ experiences were interpreted through a leadership lens. Future research could examine how faculty at different career stages experience these scholarly communities (e.g., Corral et al.^11^), or how outcomes differ across institutional contexts. Finally, while this study identifies practices associated with sustainability, it does not evaluate their relative effectiveness or cost. Such an evaluation is an important next step for institutions facing constrained resources.

## Conclusions

Sustaining scholarly communities of practice in HPE is neither straightforward nor formulaic. Our findings suggest that durable communities of practice depend on thoughtful institutional investment, intentional community cultivation, developmental patience, and visible recognition pathways, all adapted to the local context. By foregrounding the lived experiences and strategic decisions of leaders in the field, this study offers practice-informed guidance for institutions seeking to move beyond symbolic structures toward scholarly communities that are genuinely supportive, productive, and enduring.

## Data Availability

De-identified interview data collected in the present study are available upon reasonable request to the authors.

## Acknowledgments

The authors would like to sincerely thank Drs. Beth West and Tia Lockspeiser for their helpful reviews and comments on an early version of the manuscript.

## Ethical approval

Ethical approval was granted by the George Washington University Office of Human Research (letter dated December 17, 2024).

